# Dynamical SPQEIR model assesses the effectiveness of non-pharmaceutical interventions against COVID-19 epidemic outbreaks

**DOI:** 10.1101/2020.04.22.20075804

**Authors:** Daniele Proverbio, Françoise Kemp, Stefano Magni, Andreas Husch, Atte Aalto, Laurent Mombaerts, Alexander Skupin, Jorge Gonçalves, Jose Ameijeiras-Alonso, Christophe Ley

## Abstract

Against the current COVID-19 pandemic, governments worldwide have devised a variety of non-pharmaceutical interventions to suppress it, but the efficacy of distinct measures is not yet well quantified. In this paper, we propose a novel tool to achieve this quantification. In fact, this paper develops a new extended epidemic SEIR model, informed by a socio-political classification of different interventions, to assess the value of several suppression approaches. First, we inquire the conceptual effect of suppression parameters on the infection curve. Then, we illustrate the potential of our model on data from a number of countries, to perform cross-country comparisons. This gives information on the best synergies of interventions to control epidemic outbreaks while minimising impact on socio-economic needs. For instance, our results suggest that, while rapid and strong lock-down is an effective pandemic suppression measure, a combination of social distancing and contact tracing can achieve similar suppression synergistically. This quantitative understanding will support the establishment of mid- and long-term interventions, to prepare containment strategies against further outbreaks. This paper also provides an online tool that allows researchers and decision makers to interactively simulate diverse scenarios with our model.

## 1 Introduction

The current global COVID-19 epidemic has led to significant impairments of public life world-wide. To suppress the spread of the virus and to prevent dramatic situations in the healthcare systems, many countries have implemented a combination of rigorous measures like lock-down, isolation of symptomatic cases and the tracing, testing, and quarantine of their contacts. In order to gain information about the efficacy of such measures, a quantitative understanding of their impact is necessary. This can be based on statistical methods [1] and on epidemiological models [2]. While statistical methods allow for accurate characterization of the population’s health state, epidemiological modeling can provide more detailed mechanisms for the epidemic dynamics and allow investigating how epidemics will develop under different assumptions.

Preliminary efforts have been made to quantify the contribution of different policy interventions [3], but these rely on complex models based on a number of assumptions. We base our study on a classical SEIR-like epidemiological model. SEIR models are minimal mechanistic models that consider individuals transitioning through Susceptible *→* Exposed *→* Infectious *→* Removed state during the epidemics [4]. The essential control parameter is the basic reproduction number *R*_0_ [5], that worldwide non-pharmaceutical suppression strategies aim at reducing below the threshold value 1. Building on this, we incorporate additional compartments reflecting different categories of intervention strategies, identified by socio-political studies [6]. In particular, the model focuses on three main suppression programs: social distancing (lowering the rate of social contacts), active protection (decreasing the number of susceptible people), and active removal of latent asymptomatic carriers [7]. This study investigates how these programs achieve repression both individually and combined, first conceptually and then by cross-country comparison. This information can supply Government decisions, helping to avoid overloading the healthcare system and to minimise stressing the economic system (due to prolonged lock-down). We expect our model, together with its interactive online tool, to contribute to crucial tasks of decision making and to prepare containment strategies against further outbreaks.

## 2 Methods

This study links policy measures to epidemiological modelling and uses the developed model to quantitatively assess the efficacy of different interventions in six countries. In this section, we illustrate the modelling choice and the use of data.

### 2.1 The classical SEIR model

SEIR models are continuous-time, mass conservative compartment-based models of infectious diseases [4, 8]. They assume homogeneous propagation media (or fully connected graphs) and focus on the evolution of mean properties of the closed system. All of these models, from the more conceptual to more realistic versions, e.g. SEIR with delay [9], spatial coupling [10], or individual-based models [11], are classical and widely used tools to investigate the principal mechanisms governing the spread of infections and their dynamics.

Main compartments of SEIR models (see Fig. 1, framed) are: susceptible S (the pool of individuals likely to be infected), exposed E (corresponding to latent carriers of the infection), infectious I (individuals having developed the disease and being contagious) and removed R (those that have processed the disease, being either recovered or dead). The model’s default parameters are the average contact rate *β*, the inverse of mean incubation period *α* and the inverse of mean contagious period *γ*. When focusing on infection dynamics rather than patients’ fate, the latter combines recovery and death rate [12]. From these parameters, epidemiologists calculate the “basic reproduction number” *R*_0_ = *β/γ* [13] at the epidemic beginning. During the epidemic progression, isolation after diagnosis, vaccination campaigns and active suppression measures are in action. Hence, we speak of “effective reproduction number” 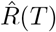 [14].

**Fig. 1.**
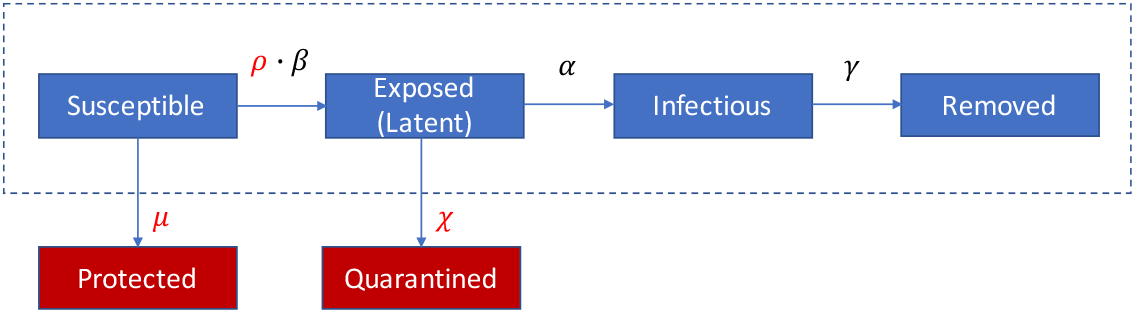
Scheme of the SPQEIR model. The basic SEIR model (framed blue blocks) is extended by the red blocks to the SPQEIR model. Parameters that are linked to repression strategies are shown in red. Interpretation and values of parameters are given in Table 2.

### 2.2 Data and analyzed countries

Governments worldwide have issued a number of social measures, including those for public health safeguard, economic support, movement restriction and non-pharmaceutical interventions to hamper disease spreading. Scholars from political sciences and sociology have recorded and classified such measures [15, 16]. Among the resources listed on the World Health Organization “Tracking Public Health and Policy Measures” [6], we used information from the ACAPS database [17] that contains a curated categorization of policy measures. ACAPS is an independent, non-profit information provider helping humanitarian actors respond more effectively to disasters. The ACAPS analysis team has aggregated and classified interventions from different sources (media, governments and international organizations), for all countries and in time. Suppression measures against the epidemic are classified under “Movement restrictions”, “Lock-down”, “Social Distancing” and “Monitoring and Surveillance”. Our modelling choice is based on these categories, which are reflected by additional compartments to the classical SEIR model (see next section).

Epidemiological data for all selected countries and regions were obtained from the COVID-19 Data Repository by the Center for Systems Science and Engineering (CSSE) at Johns Hopkins University [18]. The data are from 22 Jan 2020 to 08 July 2020. Lombardy data were obtained from the Protezione Civile Italiana data repository “Dati COVID-19 Italia” [19], from 22 Feb 2020 to 08 July 2020.

This study analyses the effect of suppression measures in flattening the curve. Despite having a precise starting date, such measures take some days to be fully effective. We estimate an average delay using the Google Mobility Reports [20, 21] for the selected countries. Google provides changes in mobility with respect to a monthly baseline, w.r.t. 6 locations: Retail & Recreation, Grocery & Pharmacy, Transit stations, Workplaces, Residential, Parks. We average the decrease in mobility at the first four locations (corresponding to those where social mixing happens more frequently [22]) to get a proxy of the time needed for hard lock-down to be fully effective (cf. Fig. 4c).

### 2.3 The extended SPQEIR model to reflect suppression strategies

SEIR models reproduce the typical bell-shaped epidemic curves for the number of infected (and still infectious) people. These quantify the main stressors for both the health system, i.e. the peak of the curve, and the economic system, i.e. the time 𝕋 passed until no new infections occur. Mainstream suppression measures against the epidemic aim at flattening the curve of new infections [7]. However, the classical SEIR model is not granular enough to investigate suppression measures when they need to be considered or should be sequentially reduced if already in place. Therefore, we extend the classical SEIR model as in Fig. 1 (red insertions) into the SPQEIR model, to reflect the intervention categories described above. The model can be summarized as follows:

- The classical blocks S, E, I, R are maintained;
- A social distancing parameter *ρ* is included to tune the contact rate *β*;
- Two new compartments are introduced where:
  − Protected P includes individuals that are isolated from the virus through lock-down, thus reducing the susceptible pool;
  − Quarantined Q describes latent carriers that are identified and quarantined after monitoring and tracing.

We do not introduce a second quarantined state for isolation of confirmed cases after the Infectious state [23, 2] but consider this together with the Removed state (see Liu et al. [24] and references therein). Quarantining infected symptomatic patients is a necessary first step in every epidemic [25]. An additional link from Q to R, even though realistic, is neglected as both compartments are already outside the “contagion system” and would therefore be redundant from the perspective of evolution of the infection. In general, protected individuals can get back to the pool of susceptible after a while, but here we neglect this transition, to focus on simulating repression programs alone. Long-term predictions could be modelled even more realistically by considering such link, that would lead to an additional parameter to be estimated and is beyond the scope of the present paper.

The model has in total 6 parameters. Three of them (*β, α, γ* introduced in Fig. 1) are based on the classical SEIR model. The new parameters *ρ, µ, χ* account for alternative repression programs (see Table 2 for details). Commonly, social distancing is modeled by the parameter *ρ*. It tunes the contact rate parameter *β*, resulting in the effective reproduction number 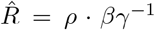. This occurs in a closed-system setting where all individuals belong to the susceptible pool, but interact less intensively with each other. The parameter *µ* stably decreases the susceptible population by introducing an active protection rate. This accounts for improvements of public health, e.g. stricter lock-down of communities, or physical reduction of a country’s population like reduced commuters’ activity. This changes the effective reproduction number into 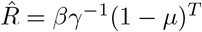 with *T* being the number of days the measures are effective [26]. The parameter *χ* introduces an active removal rate of latent carriers. Intensive contact tracing and improved methods to detect asymptomatic latent carriers may enhance the removal of exposed subjects from the infectious network. Following earlier works [27, 28] and adjusting the current parameters, 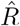 can be then expressed as 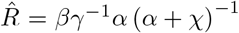. Parameter values that are not related to suppression strategies are set from COVID-19 epidemic literature [24, 29]. We use mean values as the main focus of the present model lies on sensitivity analysis of suppression parameters. Our model can be further extended by time dependent parameters [25]. Default values for suppression parameters are {*ρ, µ, χ*} = {1, 0, 0}, corresponding to the classical SEIR model.

The dynamics of our SPQEIR model is described by the following system of differential equations:

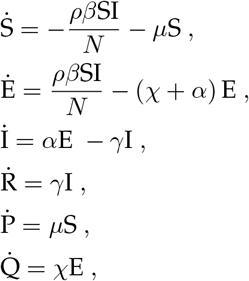

with conservation of the total number of individuals, meaning 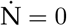 with N = S + E + I + R + P + Q. As value for the qualitative study, we used N = 10,000. For the cross-country assessment, N is adjusted to true population values for each country. Overall, the effective reproductive number becomes

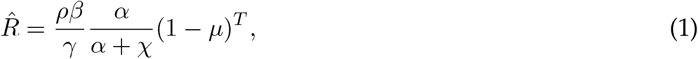

with *T* being the number of days that the measures leading to compartment P are active.

Suppression measures are initiated several days after the first infection case. Hence, we activate nondefault parameter values after a delay *τ*. In conceptual simulations, we set it arbitrarily without loss of generality. For data fitting, we fit and compare it to the official date when measures are initialized (cf. Table 1). To integrate the model numerically, we use the *odeint* function from *scipy*.*integrate* Python library.

**Table 1:** Test countries, with measures implemented (following the ACAPS database [17]), corresponding parameter in our SPQEIR model and starting date. For Lombardy, we used the Italian official date for lock-down. Differently from other countries, Ireland issued measures on two different dates; we use this case to compare social distancing and lock-down effect in a single country. We also report the (rounded) population of each country.

| Country | Measures | Param. involved | Starting Date | Population (rounded) |
| --- | --- | --- | --- | --- |
| Austria (AT) | Partial lock-down | $\mu, \rho$ | 16 Mar | 9,000,000 |
| | Social distancing | $\rho, \mu$ | 16 Mar | |
| | Contact tracing | $\chi$ | 16 Mar | |
|  | Phase-out |  | Around 14 April |  |
| Denmark (DK) | Social distancing | $\rho, \mu$ | 13 Mar | 6,000,000 |
| | Mild surveillance | $\chi$ | 13 Mar | |
|  | Phase-out |  | 14 Apr |  |
| Ireland (IR) | Partial lock-down | $\mu, \rho$ | 28 Mar | 5,000,000 |
| | Social distancing | $\rho, \mu$ | 13 Mar | |
|  | Phase-out |  | 18 May |  |
| Israel (IL) | Partial lock-down | $\mu, \rho$ | 15 Mar | 9,000,000 |
| | Social distancing | $\rho, \mu$ | 15 Mar | |
| | Contact tracing | $\chi$ | 15 Mar | |
|  | Phase-out |  | 19 April |  |
| Lombardy (LO) | Lock-down | $\mu, \rho$ | 13 Mar (Italian) | 10,000,000 |
| | Social distancing | $\rho, \mu$ | 13 Mar | |
|  | Phase-out |  | Around 15 Apr |  |
| Switzerland (CH) | Lock-down. | $\mu, \rho$ | 16 Mar | 8,500,000 |
| | Social distancing | $\rho, \mu$ | 16 Mar | |
|  | Phase-out |  | 27 Apr |  |

**Table 2:**
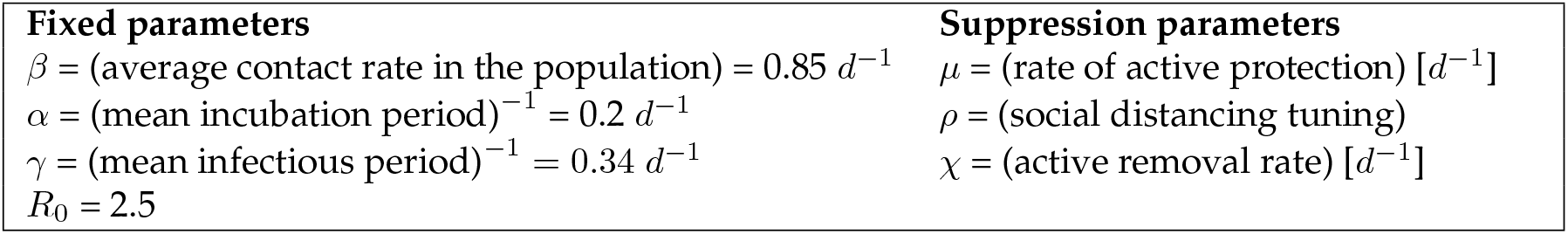
SPQEIR model parameters with their standard values for the COVID-19 pandemic from literature [24, 30]. Here “*d*” stands for days.

### 2.4 Model fitting

We fit the model to the official number of currently infected (active) cases, for each considered country. Model fitting to the infectious curves is performed in two steps, using the parameters known to be active (cf. Table 1). First, we estimate the “model consistent” date of first infection, so that the simulated curve matches the reported data of active infections. This initial step corresponds to setting the time initial conditions of the SEIR model [2, 26]. The fitting is performed with default parameter values, on a subset of data corresponding to the first outbreak, from first case until when measures are implemented (cf. Table 1). We use a grid search method for least squares, sufficient to fit a single parameter:

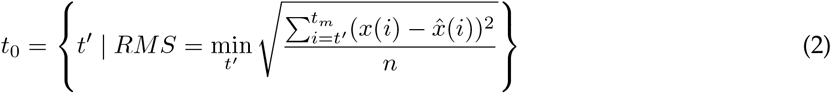

where *t*_0_ is the “model consistent” estimated date of first infection, *t*_*m*_ refers to the date measures are implemented, 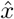 and *x* are respectively reported and model-predicted data, and *n* is the number of points between *t* and *t*_*m*_.

The second step estimates the suppression parameters that yield the best fitting of the simulated SPQEIR curve on reported data, during the first phase with implemented measures. This period is identified between the starting date *t*_*m*_ (also included in the fitting) and the phase-out date *t*_*p*_, cf. Table 1. Holding the epidemic parameters to literature values to achieve cross-country comparison on intervention parameters alone, the fitting is performed for a set of suppression parameters relative to each country, as reported by policy databases (cf. Table 1). The fit is performed with *lmfit* Python library.

When more parameters are involved at once, we also perform a comparative information analysis between our extended model and the simplest SEIR that lumps parameters under a single “social distancing” *ρ*. This shows what we gain in distinguishing the intervention parameters, not only in terms of improved interpretation, but also in terms of model fitting. We employ a reduced *χ*^2^ metric to evaluate the goodness-of-fit for both models, considering the degrees of freedom [31]:

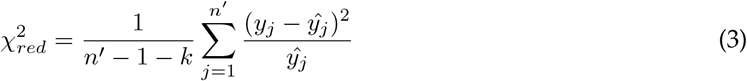

where *n* ′ is the number of data points until phase-out, *k* is the number of parameters in the model, *y*_*j*_ are estimated values and *ŷ*_*j*_ the expected ones. The lower the 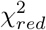 value, the better the SPQEIR model fits the data w.r.t. the simplest “social distancing” SEIR model.

## 3 Results

We first focus on the conceptual analysis of the effect of suppression interventions, initially for single measures (social distancing, active protection and active quarantining) and subsequently for a number of synergistic approaches. In particular, we study how crucial quantities, namely the infectious peak height and time to zero infectious, depend on suppression parameters and affect 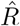. We define 𝕋 as the time when there are less than 0.5 individuals in the I compartment. This because ODE models approximate discrete quantities with continuous variables. Finally, we perform model fitting and intervention assessment over a set of countries. This provides quantitative outputs about the effectiveness of real measures, informing about the synergies applied and enabling cross comparison.

### 3.1 Mathematical analysis of single suppression measures

#### 3.1.1 Only social distancing

The parameter *ρ* captures social distancing effects, taking values in the interval [0, 1], where 0 indicates no contacts among individuals while 1 is equivalent to no actions taken. Without loss of generality, simulations consider a delay of 10 days from the first infection to the time social distancing is initiated. Fig. 2 reports simulation results. The curve of infectious is progressively flattened by social distancing (2a) and its peak suppressed (2b). However, the eradication time gets delayed for decreasing *ρ*, until a threshold yielding a disease-free equilibrium rapidly (2c). In this case, the critical value for *ρ* is 0.4, leading to 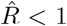. However, we notice that values of *ρ* ≃ 0.3 or lower are more effective in suppressing the epidemic faster. This is in line with early findings, suggesting that epidemic control with social distancing alone should be done “well or not at all” [32].

**Fig. 2.**
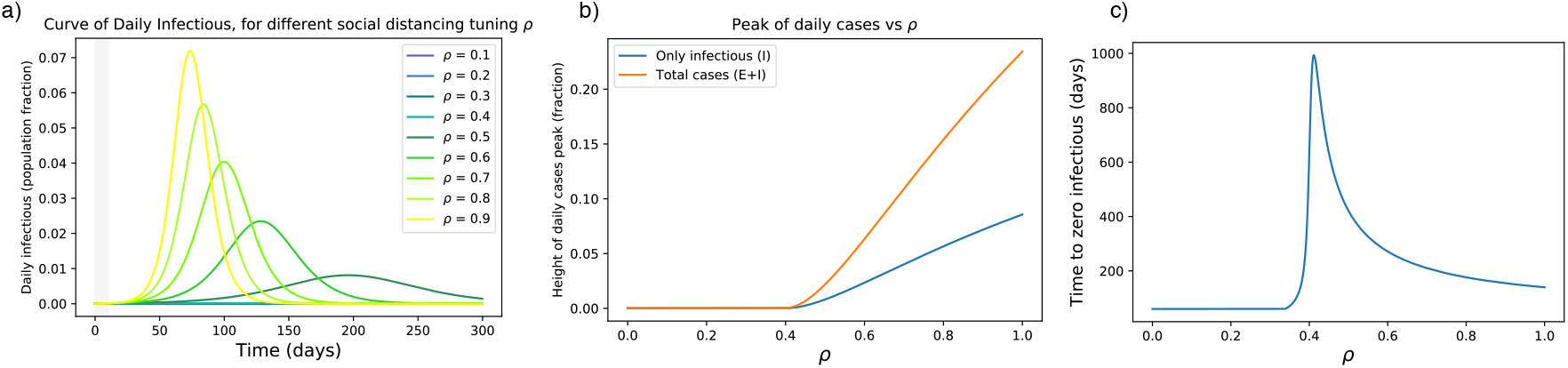
(a) Effects of social distancing on the epidemic curve. The grey area indicates when measures are not yet in place. (b) The peak is progressively flattened until a suppression is reached for sufficiently small *ρ*. For these settings, the critical value for *ρ* is 0.4 (it pushes 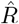 below 1). (c) Unless *ρ* is small enough, stronger measures of this kind might delay the suppression time 𝕋 of the epidemic.

#### 3.1.2 Only active protection

As above, our simulations take into account 10 days delay from the first infection to the initiation of active protection. The range of *µ* is only up to values similar to those measured in China [26]. Higher values are considered for step-wise hard lock-down (see below). The results are reported in Fig. 3. We see that small precautions can make an initial difference, but then the effects saturate (Fig. 3a,b). The time to zero infectious is decreased with higher values of active protection (Fig. 3a,b). In particular, *µ* = 0.01 *d*^*−*1^ suppresses the epidemics in about 6 months by protecting 70% of the population. Higher values of *µ* achieve suppression faster, while protecting almost 100% of the population. If protection is mostly achieved through isolation, this is unrealistic. However, this models effective vaccination strategies.

**Fig. 3.**
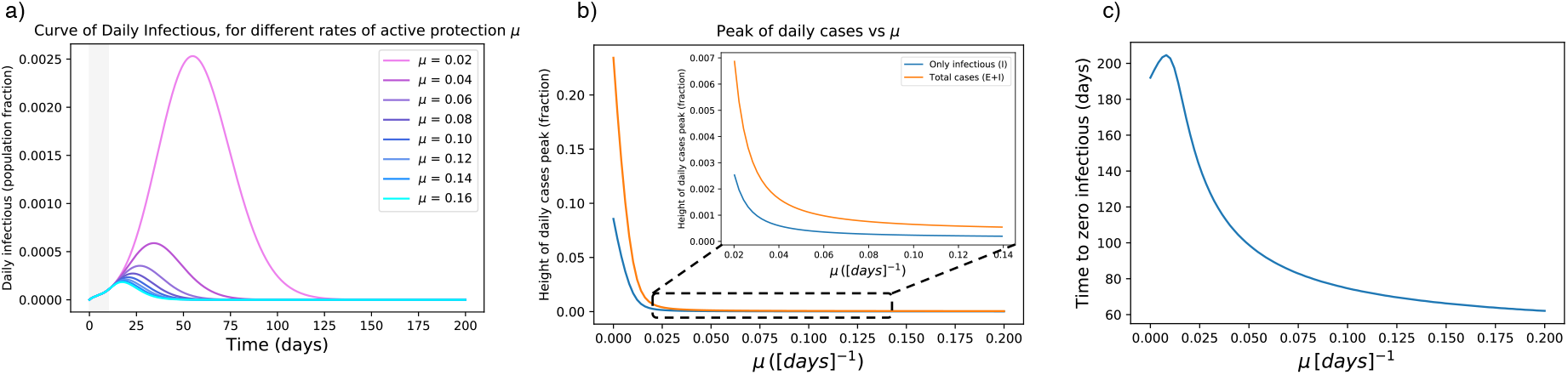
(a) Effects of active protection on the infectious curve. The grey area indicates when measures are not yet in place. *µ* is expressed in *d*^*−*1^. (b) Dependency of peak height on *µ*: the peak is rapidly flattened for increasing *µ*, then it is smoothly reduced for higher parameter values. (c) High *µ* values are effective in anticipating the complete eradication of the epidemic, but require protecting more than 90% of the population.

**Fig. 4.**
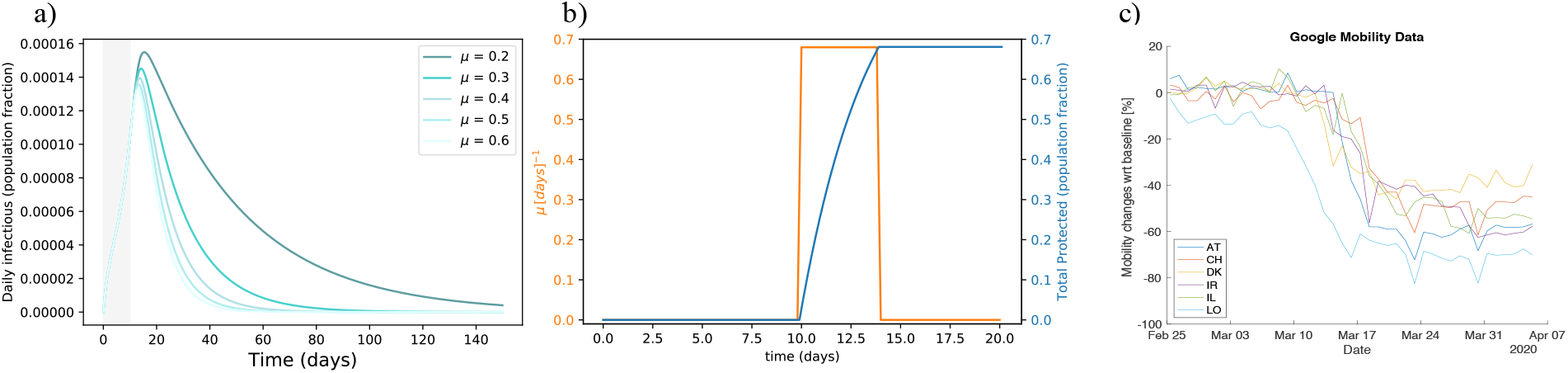
(a) Flattening the infectious curve by hard lock-down. Rapidly isolating a large population fraction is effective in suppressing the epidemic spreading. (b) Modeling hard lock-down: high *µ*_*ld*_ (orange) is active for four days to isolate and protect a large population fraction rapidly (blue). As an example, we show *µ*_*ld*_ = 0.28 *d*^*−*1^ if *t* ∈ [10, 14]. It results in protecting about 68% of the population in two days. Higher values, e.g. *µ*_*ld*_ = 0.65 *d*^*−*1^ would protect 93% of the population at once. (c) Google Mobility Report visualization [21] for analysed countries, around the date of measures setting. Each line reports the mean in mobility change across Retail & Recreation, Grocery & Pharmacy, Transit stations, and Workplaces, around the date of implementation of the measures. A minimum of 4 days (from top to bottom of steep decrease) is required for measures to be fully effective. Abbreviation explanations: AT = Austria, CH = Switzerland, DK = Denmark, IL = Israel, IR = Ireland, LO = Lombardy.

We also consider hard lock-down strategies which isolate many people at once [33]. This corresponds to reducing S to a relatively small fraction rapidly. Since *µ* is a rate, we mimic a step-wise hard lockdown by setting a high value to *µ*, but its effect only lasts for a short period of time, see Fig. 4b. We thus use the notation *µ*_*ld*_. In the figure, an example shows how to rapidly protect about 68% of the population with a step-wise *µ*_*ld*_ function. In particular, we use an average four-days long step-wise *µ*_*ld*_ function (Fig. 4b) to mimic the rapid, but not abrupt, change in mobility observed in many countries by Google Mobility Reports [21] (Fig. 4c). Lock-down effects are reported in Fig. 4a: a hard lock-down is effective in suppressing the epidemic curve and in lowering the eradication time.

#### 3.1.3 Only active quarantining

The simulations in this part are based on realistic assumptions: testing a person is effective only after a few days that that person has been exposed (to have a viral charge that is detectable). This induces a maximal quarantining rate *θ*, which we set *θ* = 0.33 *d*^*−*1^ as testing is often considered effective after about 3 days from contagion [34]. Therefore, we get the active quarantining rate *χ* = *χ*′ · *θ*, where *χ* ′ is a tuning parameter associated e.g. to contact tracing. As *θ* is fixed, we focus our analysis on *χ* ′. As above, we also assume that testing starts after the epidemic is seen in the population, i.e. some infectious are identified with the usual 10 days delay in the activation of measures.

The corresponding results are reported in Fig. 5. The curve is progressively flattened by latent carriers quarantining and its peak suppressed, but the eradication time gets delayed for increasing *χ* ′. This happens until a threshold value of 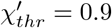 that pushes 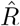 below 1. This value holds if we accept a strategy based on testing, with *θ* = 0.33. If preventive quarantine of suspected cases does not need testing (for instance, it is achieved by contact tracing apps), the critical *χ* ′ value could be drastically lower. In particular, 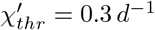, if *θ* = 1*d*^−1^, i.e. latent carriers are quarantined the day after a contact.

**Fig. 5.**
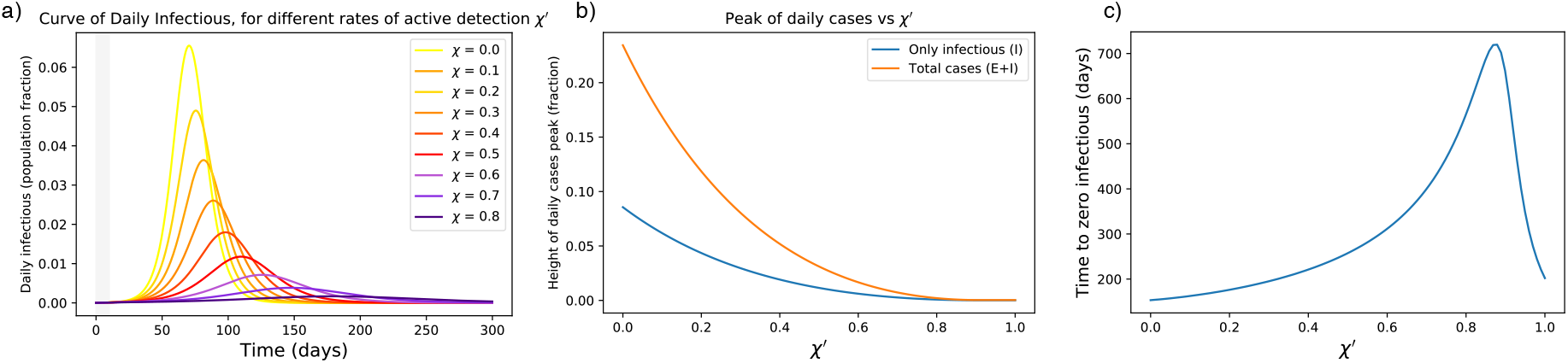
(a) Effects of active latent carriers quarantining on the epidemic curve. The grey area indicates when measures are not yet in place. (b) The peak is progressively flattened until a disease-free equilibrium is reached for sufficiently large *χ*. (c) Unless *χ* ′ is large enough, stronger measures of this kind might delay the complete eradication of the epidemic. Note that the critical *χ* ′ can be lowered for higher *θ*, e.g. if preventive quarantine does not wait for a positive test.

The parameter *χ* ′ tunes the rate of removing latent carriers. Hence, it combines tracing and testing capacities, i.e. probability of finding latent carriers (*P*_*find*_) and probability that their tests are positive (*P*_+_). The latter depends on the false negative rate *δ*_*−*_ as

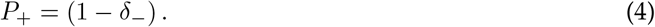

So, *χ* ′ = *P*_*find*_ · *P*_+_. Hence, suppressing the peak of infectious requires an adequate balance of accurate tests and good tracing success as reported in Fig. 6. Further quantifying the latter would drastically improve our understanding of the current capabilities and of bottlenecks, towards a more comprehensive feasibility analysis.

**Fig. 6.**
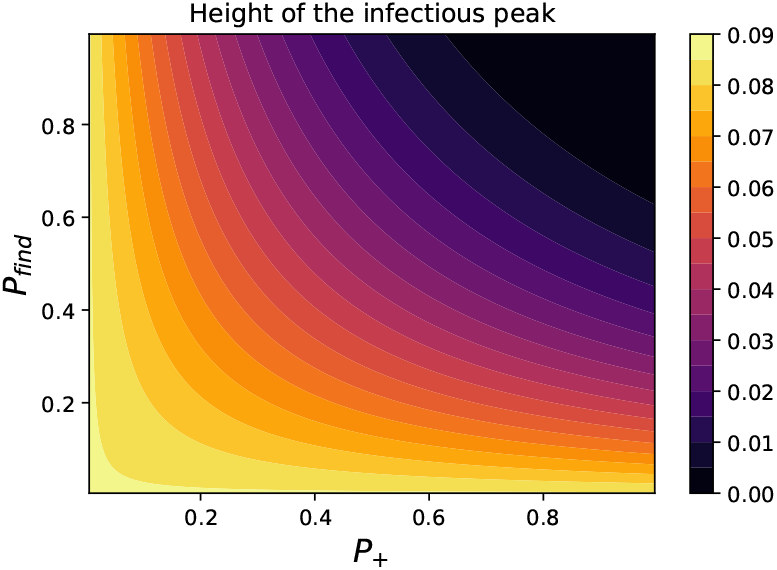
Assessing the impact of *P*_*find*_ and *P*_+_ on the peak of infectious separately. This way, we separate the contribution of those factors to look at resources needed from different fields, e.g. network engineering or wet lab biology. Solutions to boost the testing capacity like [35] could impact both terms.

### 3.2 Synergistic scenarios

Fully enhanced active quarantining and active protection might not be always feasible, e.g. because of limited resources, technological limitations or welfare restrictions. Therefore a synergistic approach is very attractive as it can flatten the curve. This section shows a number of possible synergies, concentrating as before on abstract scenarios to investigate the effect of combining different suppression programs. As case studies, we consider the 6 synergistic scenarios listed below. Parameters are set without being specific to real measures taken: their value is so far conceptual and meaningful when compared across scenarios. Just like above, we consider a 10 days delay from the first infection to issuing measures; as suggested in other studies [36], delaying action could worsen the situation. To differentiate between a rapid isolation and a constant protection, we introduce *µ*_*ld*_ (for “hard lock-down strategies”, see Section 3.1.2) separated from *µ*. To get 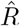, we follow Eq. 1, considering *χ* = *χ* ′ · *θ* as in Section 3.1.3 and *T* = 4 (along the steep decay after measures are in place, in a best case scenario). Our scenarios are the following:

1. Many European countries opted for a lock-down strategy. A quite large fraction of the population was isolated, individuals were recommended to self-quarantine in case of suspected positiveness, social distancing got mandatory but was sometimes not fully followed, masks and sprays were suggested for protection. So, we set an initial “hard lock-down” *µ*_*ld*_ = 0.12 *d*^*−*1^ to protect around 38% of the population quickly. Then we chose *ρ* = 0.7, *χ* ′ = 0.15 and *µ* = 0.008 *d*^*−*1^. This yields 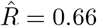.
2. An alternative procedure is to rapidly protect only the population fraction at high risk (*µ*_*ld*_ = 0.06 *d*^*−*1^, driving 15% of initial S to P). Then, we assume an improvement in individual safety giving *µ* = 0.01 *d*^*−*1^. Social distancing is relaxed (*ρ* = 0.8) but latent carrier quarantine is enforced (*χ* ′ = 0.5). This gives 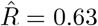.
3. In case preventive quarantine of latent carriers is not greatly effective (*χ* ′ = 0.1), and in case of low protection rate and scarce isolation (*µ* = 0.004 *d*^*−*1^, *µ*_*ld*_ = 0.08 *d*^*−*1^), we rise social distancing for all individuals doing business as usual (*ρ* = 0.5). In this case, 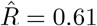.
4. If there are no safety devices that provide an adequate protection (*µ* = 0 *d*^*−*1^), we set *ρ* = 0.45, *µ*_*ld*_ = 0.2 *d*^*−*1^ and *χ* ′ = 0.2 to get 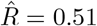.
5. This case has higher 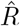 than the previous ones, namely 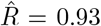. The corresponding parameters are *µ*_*ld*_ = 0.1 *d*^*−*1^, *µ* = 0.002 *d*^*−*1^, *ρ* = 0.7, *χ* ′ = 0.1.
6. Finally, we consider “draconian” measures such that 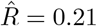 only through isolation and massive latent carriers quarantining. So, *µ*_*ld*_ = 0.6 *d*^*−*1^ and *χ* ′ = 0.3 while *ρ* = 1 and *µ* = 0 *d*^*−*1^.

Simulation results are reported in Fig. 7. Different synergies lead to different timing, even though the peak is contained similarly (Fig. 7a). This has an impact on the cumulative number of cases (Fig. 7b) that will be reflected on the death toll. This holds even when the 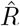 values are very close, as in scenarios 1 to 4. Focusing on scenarios 2 and 3, we notice that prevention measures and latent quarantine accelerate the suppression, even when isolating only vulnerable people. This achieves similar effects as strong social distancing. In addition, active protective measures with relatively low values further concur in suppressing the peak. This finding asks for rapid assessment of masks and sanitising routines.

**Fig. 7.**
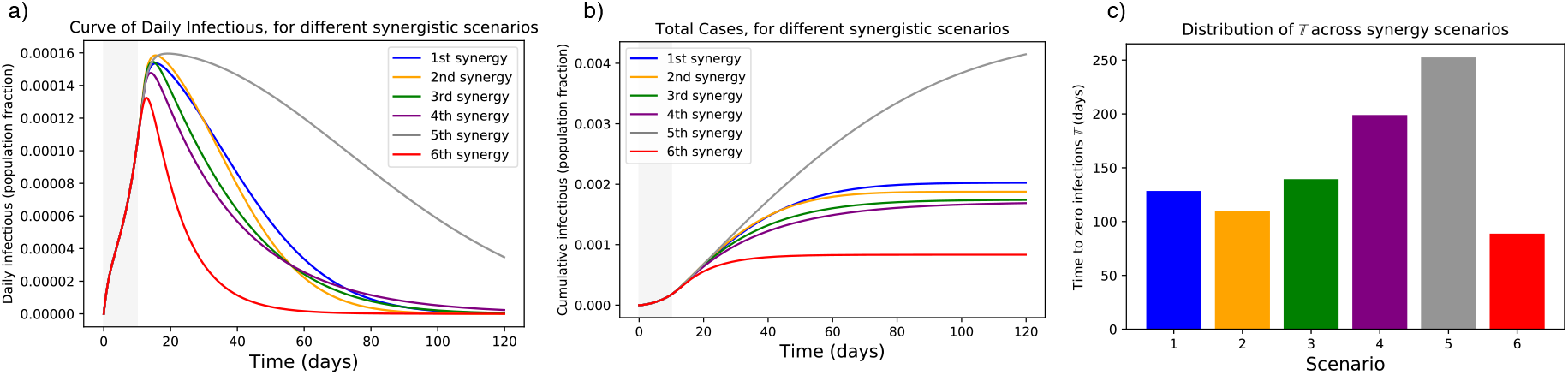
Simulations of the 6 synergistic scenarios. (a) Curves of infectious Individuals, (b) Cumulative cases. The grey area indicates when measures are not yet in place. It is evident that scenarios leading to the same 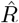 could show different patterns and suppression timing. (c) Distribution of times to zero infections 𝕋 for different scenarios.

Overall, the strength of suppression measures influences how and how fast the epidemic is flattened. *µ*_*ld*_ mostly governs the peak height after measures are implemented, *ρ* mainly tunes the curve steepness together with *µ*, while *χ* shifts the decaying slope up and down. Overall, a 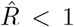 suffices to avoid breakdown of the health system, but its effects could be too slow for the economic system. Decreasing its value with synergistic interventions could speed up epidemic suppression. A careful assessment of measures’ strength is thus recommended for cross-country comparison.

### 3.3 Model fitting and interventions assessment

#### 3.3.1 Model fitting

As described in the Methods section, we first estimate the “model consistent” date of first infection *t*_0_, that is, the temporal initial condition for the SPQEIR model. We do not claim that this is the true date of first infection in a country. On the contrary, it is the starting date of infections in case of homogeneous transmission, under the assumption of no superspreading events [37], and with the hypothesis of coherent *R*_0_ (cf. Table 2). During the second fitting step, we also estimate the date at which suppression measures start having effect on the infectious curve, *t*_*m*_. Comparing *t*_*m*_ with official intervention dates from Table 1, we notice that about 8 days are necessary to register lock-down effects. This is consistent to early findings on lock-down effectiveness [38]. Estimated dates are reported in Table 3.

**Table 3:** Dates of official detection of first COVID-19 case [18], estimated dates for first infection *t*_0_ (according to Eq. 2) and date at which measures start being effective *t*_*m*_, per country. Although consistent with recent literature [26] that suggests the first infection happened weeks before the first official detection, these retrospective dates should be interpreted under the model assumptions.

| Country | AT | DK | IR | IL | LO | CH |
| --- | --- | --- | --- | --- | --- | --- |
| 1<sup>st</sup> official detection | 24 Feb | 04 Mar | 29 Feb | 21 Feb | 21 Feb | 25 Feb |
| $t_0$ | 22 Jan | 22 Jan | 29 Jan | 24 Jan | 05 Jan | 14 Jan |
| $t_m$ | 26 Mar | 21 Mar | 06 Apr | 30 Mar | 19 Mar | 21 Mar |

Then, we fit suppression parameters to data affected by policy measures, from their estimated starting date *t*_*m*_ to phase-out *t*_*p*_ (cf. Table 1). Results of the model fitting are reported in Fig. 8. The SPQEIR model, with appropriate parameters for each country, is fitted to reported infection curves and, overall, model fitting have good agreement with data. This supports the model structure as very simple yet realistic enough to capture the main dynamical behaviour of the infection curves in multiple countries. In addition, it allows for each country to obtain multiple sets of parameters, providing a good fit and representing different strategies. Finally, it allows a comparison between different countries through the corresponding best fit parameters. For Ireland, although initial social distancing advises were issued on 13th March (cf. Table 1), fitting the complete curve was only possible when considering the lock-down date (28th March) as the major driver of the suppression.

**Fig. 8.**
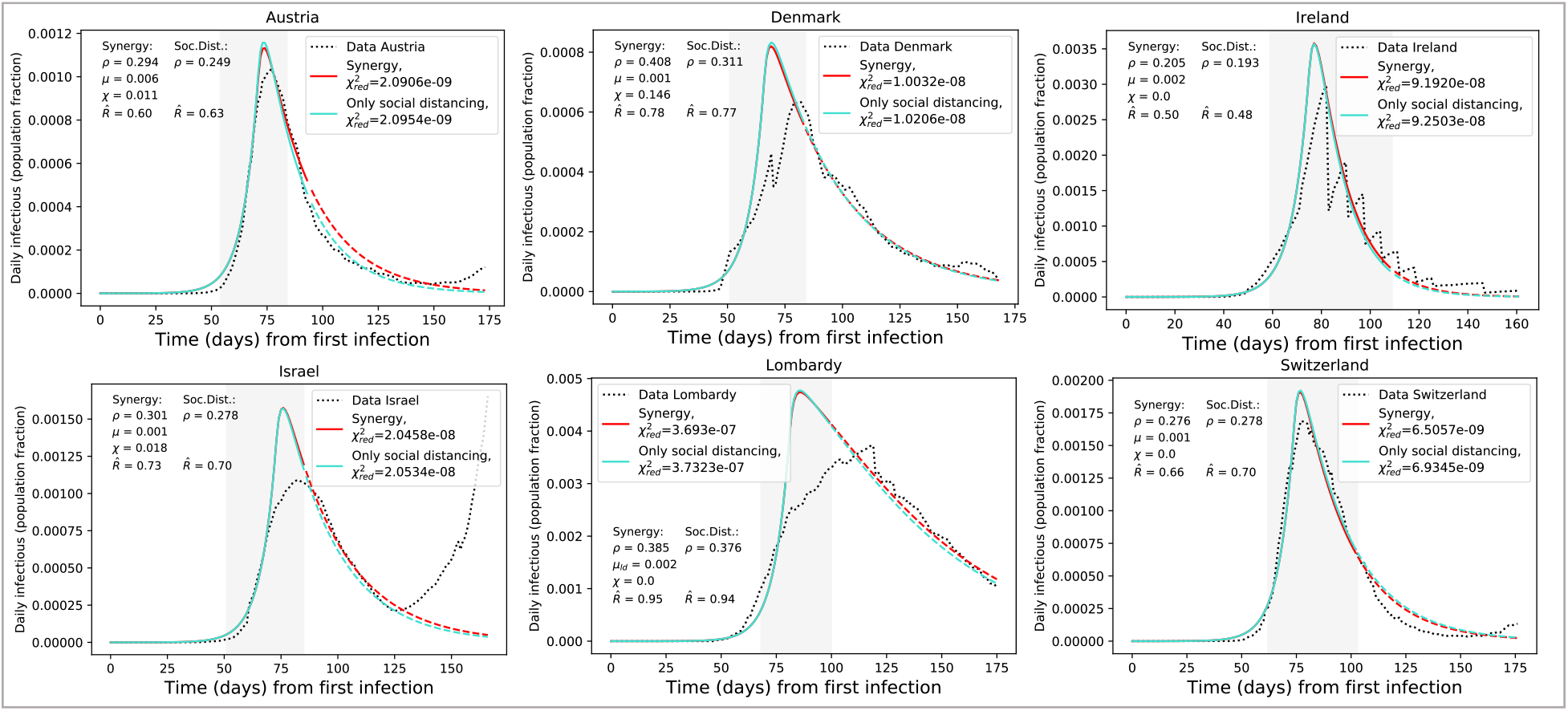
Results of model fitting. Infection curves for the considered countries (dotted) are fitted with the SPQEIR model with appropriate parameters (red curves). We also show a comparison with the fitted curve obtained from the “basic” SEIR model with only social distancing (turquoise curves). Parameter values are reported for each country, as well as the corresponding 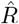 (for the gray area, following Eq. 1) and 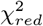. The period of measures enforcement, from *t*_*m*_ to *t*_*p*_, is highlighted by the grey region. Time progresses from the estimated day of first infection *t*_0_ (cf. Table 3). Population fraction refers to country-specific populations (cf. Table 1). After phase-out, we prolong the fitted curve (parameter values unchanged) to compare observed data with what could have been if measures had not been lifted (dashed lines). From the data, we can observe a resurgence of cases that points to possible “second outbreaks” (particularly in Israel).

The model matches very well those regions like Austria and Switzerland that avoided saturation of the healthcare system and thus reported reliable data (cf. Fig. 4c). Ireland reported intermittent data, while Lombardy is not perfectly represented, probably because of data reporting and larger heterogeneity in its spacial patterns.

Finally, the reduced *χ*^2^ metric (Eq. 3) usually attains minimum values for the complete SPQEIR model rather than the simple social distancing one. Hence, the SPQEIR is confirmed to be informative, on top of being fully interpretable and linked to recognised social policy categories.

#### 3.3.2 Cross-country interventions assessment

Fitting a number of countries with the same model containing the same epidemiological parameters allows to perform a quantitative and consistent comparison on the efficacy of their interventions. In Fig. 8, parameter values providing the best fit of model to data are reported, together with the simulation results (mean values) calculated by *lmfit* algorithm. Different synergies yield similar values for 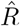, but the curve is different in its evolution as already observed in the previous sections. As expected from the model analysis above, the lower 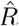 is (below 1), the faster the suppression of the epidemic. In addition, different parameter combinations generate differing curves, which might well explain differences in reported total cases and deaths between various countries. Comparing Austria, Denmark and Lombardy we can see that contact tracing and monitoring might play a role in speeding up the curve decay, despite the fact that population-wide interventions played a major role. In general, combined isolation and tracing strategies would reduce transmission in addition to social distancing or self-isolation alone. In particular, social distancing alone is effective only if very stringent, as suggested by “only social distancing” fitting. In general, a strong, rapid lock-down seems the best option, as also suggested by the conceptual analysis. However, intervening with additional synergies is a viable option to suppress the epidemic faster and with lower lock-down values.

Finally, we observe the value of timely interventions: we see that intervening earlier with respect to the date of first infection helps reducing the daily curves by almost a factor of 10. For instance, we can compare Denmark and Lombardy in Fig. 8: the first one got a peak corresponding to about 0.08% of the whole population, while the second region registered a number of active cases of about 0.5% of the whole population. This translates in more than 3800 infectious on the Danish peak, and on more than 37000 on Lombardy’s.

## 4 Discussion

The model is fitted until phase-out dates, when measures are progressively lifted and therefore the model assumptions do not hold anymore. In Fig. 8 we extrapolate the model, with same parameter values, after phase-out (dashed lines), to compare observed data to the most optimistic scenario, where measures would not have been lifted. We observe that, up to July 8th, the infection curves mostly maintained an inertial decreasing trend: despite some fluctuations that make them generally higher than the best scenario, they kept on following a downward trend similar to that of the model. We speculate that this phenomenon is linked to changed behaviors, face masks [39] and improved sanitising practices that maintained social distancing values, as well as contact tracing practices issued by many countries along with the phase-out. However, some countries (Israel in particular, but also Austria) showed a worrisome upward trend, possibly associate to a second outbreak. As this is not a low probability event, we stress the usefulness of our analysis to prepare for future developments in pandemic progression. It has been asked whether the peak of infections was reached because of herd immunity or because of interventions [40]. An added value of this study is to confirm that the peak of infection, for the considered countries, was not reached because of herd immunity. On the contrary, it is the effect of a number of suppression measures that reduced the number of cases artificially. This should warn about the high numbers of people that are still susceptible.

We acknowledge the limitations of our analysis. Due to its structure and the use of ordinary differential equations, the model only accounts for average trends. However, it cannot reproduce fluctuations in the data, being them intrinsic in the epidemic, or from testing and reporting protocols that might differ among countries. In addition, the constant nature of parameters used in this analysis allows good agreement between model and data when countries implemented rapid and strong measures point-wise in time, with little follow-ups. Further statistical studies, with time varying parameters, could obtain more precise values. In the same way, transferring models from country to country requires fulfilling the same assumptions on model structure and basic hypothesis. This is shown by the different fitting performances, that suggest that a transfer is not always possible.

In general, this study is not intended to make a ranking of country responses, nor to suggest that different strategies could have led to better outcomes. Contrariwise, it should be used as a methodological step towards quantitatively inquiring the effect of different intervention categories. It examines possible abstract scenarios and compares quantitative, model-based outputs, but it is not intended to fully represent specific countries nor to reproduce the epidemic complexity within societies. In fact, the model does not provide fine-grained quantification of specific interventions, e.g. how effective masks are in protecting people, how much proximity tracing apps increase *P*_*find*_, how changes in behavior are associated with epidemic decline [41] and so on. We acknowledge that the new compartments cannot perfectly match policy measures, but are a reasonable approximation. Some real measures might also affect multiple parameters at once, e.g. safety devices and lock-down could impact both *µ* and *ρ*. Comparing results of this macro-scale model with those of complex, micro-scale ones [3] could inform researchers and policy makers about the epidemic dynamics and effective synergies to hamper it. Any conclusion should be carefully interpreted by experts, and the feasibility of tested scenarios should be discussed before reaching consensus.

## 5 Conclusion

We have developed a minimal model to link intervention categories against epidemic spread to epidemiological model compartments. This allows quantitative assessment of non-pharmaceutical suppression strategies on top of social distancing, for a number of countries. Strategies have different effects on epidemic evolution in terms of curve flattening and eradication timing. As with previous studies [22, 42], we have observed the need to enforce containment measures (i.e., detect and isolate cases, identify and quarantine contacts and at risk neighborhoods) along with mitigation (i.e., slow down viral spread in the community with social distancing).

By extending the classic SEIR model into the SPQEIR model, we distinguished the impact of different suppression programs in flattening the peak and anticipating the eradication of the epidemic. Depending on their strength and synergy, non-pharmaceutical interventions can hamper the disease from spreading in a population. First, we performed a complete sensitivity analysis of their effects, both alone and in synergy scenarios. Then, we moved from idealised representations to fitting realistic contexts, allowing preliminary mapping of intervention categories to abstract programs. We verified that the model is informative in interpolating the infection curves for a number of countries, and performed cross-country comparison. We could then obtain model-based outputs on the strength of interventions, for a number of countries that respected the model assumptions. This provides better, quantitative insights on the effect of suppression measures and their timing, and allows improved comparison.

Overall, this work could contribute to quantitative assessments of epidemic suppression strategies. To tackle current epidemic waves, and against possible resurgence of contagion [43] (also cf. Fig 8), better understanding the effect of different non-pharmaceutical interventions could help planning mid- and long-term measures and to prepare preventive plans, until a vaccine is available.

## 6 Shinyapp

A user-friendly online shinyapp to interactively simulate different scenarios with the SPQEIR model is available on: https://jose-ameijeiras.shinyapps.io/SPQEIR_model/. It allows to reproduce the present outputs and to perform sensitivity analysis.

## Data Availability

No data are used. Simulation code is publicly available.

https://github.com/daniele-proverbio/Covid-19

## Ethics

The application of anonymized data for the purpose of epidemic modelling has been endorsed by the accessed databases.

## Data accessibility

Databases of social measures can be accessed at https://www.who.int/emergencies/diseases/novel-coronavirus-2019/phsm [6].

ACAPS database is at https://www.acaps.org/covid19-government-measures-dataset [17]. Worldwide epidemiological data collection from John Hopkins University is at https://github.com/CSSEGISandData/COVID-19 [18].

Lombardy data were retrived from https://github.com/pcm-dpc/COVID-19 [19].

Google mobility data [21] were accessed through https://ourworldindata.org/covid-mobility-trends [20].

The code for analysis can be found at https://github.com/daniele-proverbio/assessing_strategies.

## Authors’ Contributions

DP, AH designed the study. DP, FK, SM developed the model. DP, FK, JAA implemented the model. DP, FK, SM, AH, AA, LM, AS, JG, CL analyzed and interpreted the results. SM, AH, AS, JG, CL supervised and coordinated the project. DP, FK, SM, CL wrote the first draft. All authors contributed to the final draft. All authors gave their final approval for publication.

## Acknowledgments

The authors thank the Research Luxembourg - COVID-19 Taskforce for mutual collaborations.

